# Plasma proteomic comparisons change as coverage expands for SomaLogic and Olink

**DOI:** 10.1101/2024.07.11.24310161

**Authors:** Mary R. Rooney, Jingsha Chen, Christie M. Ballantyne, Ron C. Hoogeveen, Eric Boerwinkle, Bing Yu, Keenan A. Walker, Pascal Schlosser, Elizabeth Selvin, Nilanjan Chatterjee, David Couper, Morgan E. Grams, Josef Coresh

## Abstract

The number of assays on highly-multiplexed proteomic platforms has grown ten-fold over the past 15 years from less than 1,000 to >11,000. The leading aptamer-based and antibody-based platforms have different strengths. For example, Eldjarn et al^1^ demonstrated that the aptamer-based SomaScan 5k (4,907 assays, assessed in the Icelandic 36K) and the antibody-based Olink Explore 3072 (2,931 assays, assessed in the UK BioBank) had a similar number of *cis*-pQTLs among all targets (2,120 vs. 2,101) but Olink had a greater number of *cis*-pQTLs among the overlapping targets (1,164 vs. 1,467). Analysis of split plasma measures showed the SomaScan assays to be more precise: median coefficient of variation (CV) of 9.9% vs. 16.5% for Olink.^1^ Precision of the newest versions of the platforms—SomaScan 11k (>11,000 assays, released in December 2023) and Olink Explore HT (>5,400 assays, released in July 2023)—has not yet been established. We assessed the reproducibility of the SomaScan 11k and Olink Explore HT using split plasma samples from 102 Atherosclerosis Risk in Communities (ARIC) Study participants. We found that the SomaScan 11k assays had a median CV of 6.8% (vs 6.6% for the subset of assays available on the SomaScan 5k) and the Olink Explore HT assays had a median CV of 35.7% (vs 19.8% for the subset of assays available on the Olink Explore 3072). Across Olink assays, the CVs were strongly negatively correlated with protein detectability, i.e., percent of samples above the limit of detection (LOD). For the 4,443 overlapping assays, the distribution of between-platform correlations was bimodal with a peak at *r*∼0 and with another smaller peak at *r*∼0.8. These findings on precision are consistent with the updated results by Eldjarn et al^1^ but indicate that precision of these two leading platforms in human plasma has diverged as the number of included proteins has increased.

---

Using data from the Atherosclerosis Risk in Communities (ARIC) Study, we provide estimates of precision based on split plasma samples for the SomaScan 11k and for the Olink Explore HT, and report on the cross-platform agreement for proteins overlapping on the SomaScan 11k and Olink Explore HT. Plasma samples from 116 participants were assayed, of which 102 had measurements on both the SomaScan 11k and Olink Explore HT platforms. The 102 participants were mean aged 74 years (SD 5 years), 47% self-identified their race as Black, and 53% were women (**Extended Data Table 1**). The mean eGFR was 62 mL/min/1.73m^2^ (SD 21), 38% had diabetes, and 78% had hypertension. By design, none of the participants had cardiovascular disease at the time of blood draw, and half of the participants developed incident cardiovascular disease within the 5 years following visit 5.

The median Spearman correlation for the 11,083 aptamers (9,685 unique proteins based on uniprotID) measured in 102 split samples was 0.85 (interquartile interval [IQI]: 0.70-0.94), and the median CV was 6.8% (IQI: 5.1-9.4%; CV<20% for 10,478 assays) (**Figure 1A and Table 1**). Assay precision was similar when we stratified aptamers by their prior availability on the SomaScan 5k and 7k platforms (**Table 1**).

**Table 1.** Summary of within-platform precision (overall and according to availability on prior versions) among 102 ARIC visit 5 participants.

|  | N participants | N proteins | Summary of Spearman's *r* |  |  |  | Summary of CV |  |  |  |
| --- | --- | --- | --- | --- | --- | --- | --- | --- | --- | --- |
|  |  |  | Mean | 25<sup>th</sup> %ile | 50<sup>th</sup> %ile | 75<sup>th</sup> %ile | Mean | 25<sup>th</sup> %ile | 50<sup>th</sup> %ile | 75<sup>th</sup> %ile |
| **SomaScan 11k** | 102 | 11,083 | 0.79 | 0.70 | 0.85 | 0.94 | 8.7% | 5.1% | 6.8% | 9.4% |
| Proteins in SomaScan 11k but not 5k, 7k |  | 3,487 | 0.74 | 0.62 | 0.77 | 0.90 | 9.0% | 5.2% | 7.1% | 10.2% |
| Proteins in SomaScan 7k but not 5k |  | 2,312 | 0.82 | 0.73 | 0.87 | 0.95 | 8.6% | 5.1% | 6.8% | 9.2% |
| Proteins in SomaScan 5k, 7k, and 11k |  | 5,284 | 0.82 | 0.74 | 0.87 | 0.95 | 8.5% | 5.0% | 6.6% | 8.9% |
| **Olink Explore HT (5k)** | 102 | 5,420 | 0.61 | 0.37 | 0.65 | 0.90 | 110% | 14.5% | 35.7% | 97.1% |
| Proteins in Olink 5k but not 96, 3k |  | 2,591 | 0.47 | 0.28 | 0.45 | 0.66 | 173% | 35.4% | 67.9% | 203% |
| Proteins in Olink 3k but not Olink 96* |  | 2,394 | 0.73 | 0.55 | 0.84 | 0.94 | 56.5% | 10.8% | 19.8% | 44.2% |
| Proteins in Olink 96, 3k, and 5k |  | 435 | 0.85 | 0.83 | 0.92 | 0.96 | 26.6% | 7.6% | 10.9% | 18.3% |
| **Olink Explore HT (5k, impute <LOD to LOD/2)\*\*** | 102 | 5,160 | 0.72 | 0.58 | 0.79 | 0.93 | 24.4% | 8.7% | 13.3% | 21.7% |
| Proteins in Olink 5k but not 96, 3k |  | 2,410 | 0.64 | 0.48 | 0.70 | 0.84 | 31.0% | 9.0% | 14.7% | 24.6% |
| Proteins in Olink 3k but not Olink 96* |  | 2,317 | 0.78 | 0.68 | 0.86 | 0.95 | 19.4% | 8.9% | 12.7% | 20.3% |
| Proteins in Olink 96, 3k, and 5k |  | 433 | 0.87 | 0.84 | 0.93 | 0.96 | 14.1% | 7.3% | 10.1% | 15.4% |
| **Olink Explore HT (5k, excluding values <LOD) †** | 102 | 4,743 | 0.75 | 0.68 | 0.88 | 0.96 | 31.0% | 11.2% | 17.6% | 28.5% |
| Proteins in Olink 5k but not 96, 3k |  | 2,111 | 0.68 | 0.57 | 0.81 | 0.95 | 43.6% | 15.8% | 23.6% | 36.9% |
| Proteins in Olink 3k but not Olink 96* |  | 2,208 | 0.80 | 0.75 | 0.89 | 0.95 | 22.0% | 9.9% | 14.5% | 22.6% |
| Proteins in Olink 96, 3k, and 5k |  | 424 | 0.86 | 0.85 | 0.93 | 0.96 | 14.7% | 7.5% | 10.4% | 15.6% |

Abbreviations: ARIC, Atherosclerosis Risk in Communities; LOD, limit of detection; CV, coefficient of variation Bland-Altman.
SomaScan 11k results are ANML SMP and Olink Explore HT (5k) results are plate-normalized. Of the SomaScan platforms listed, SomaScan 5k was released first, followed by SomaScan 7k, and then SomaScan 11k. Of the Olink platforms listed, Olink 96 was released first, followed by Olink 3072 (3k), and then Olink Explore HT (5k).
\* The CVD 2, 3, inflammation, organ damage and cardiometabolic panels data (460 proteins) were included as part of the Olink 96 platform.
\*\* Impute NPX values reported below the LOD to half of the LOD (we excluded 260 proteins where all samples had Normalized Protein eXpression (NPX) values below the LOD in either batch or proteins that were missing an LOD).
† We excluded 677 proteins where <2 samples had a NPX value that was above the LOD in both batches or proteins that were missing an LOD. The median percentage of observations above the LOD were 18.6% overall and 5.9% in proteins in Olink 5k but not 96, 3k, 96.1% proteins in Olink 3k but not Olink 96, and 100% in proteins in Olink 96, 3k, and 5k

**Figure 1.**
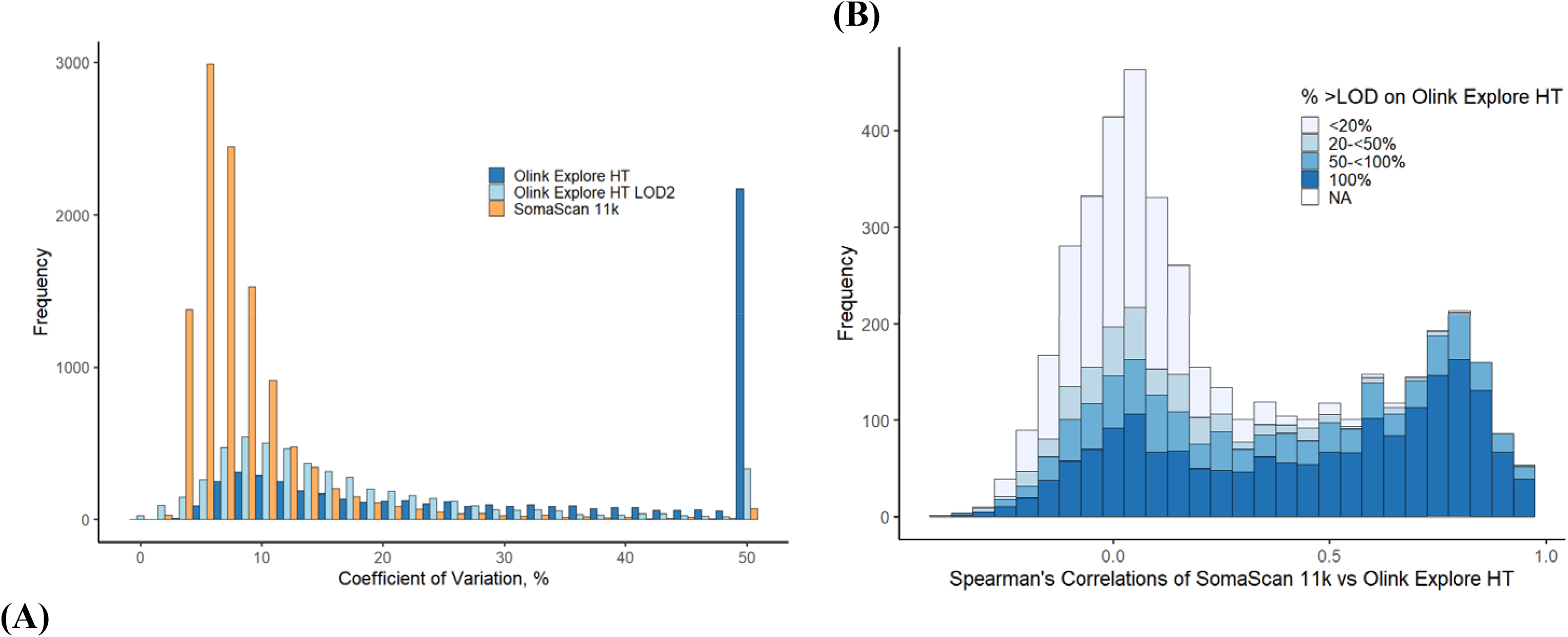
A-B. Summary of within-platform precision and between-platform correlations among 102 ARIC Study participants (2011-2013). <u>Panel A:</u> Precision of proteins measured in duplicate on the SomaScan 11k (11,083 assays) and on the Olink Explore HT (5,420 assays) before and after imputing values below the limit of detection (LOD) to the LOD divided by 2 (LOD2). Coefficients of variation (CV) were capped at 50% for the display of the histogram. <u>Panel B:</u> Histogram of Spearman’s correlations for the 4,433 overlapping protein assay comparisons on the SomaScan 11k and Olink Explore HT platforms, according to the percentage of samples with protein values above the Olink Explore HT LOD. There were 1,831 overlapping proteins with 100% samples with protein values above the LOD (i.e. detectable), 804 proteins with 50-<100%, 407 proteins with 20-<50%, 1,399 with less than 20% above the LOD; two proteins were missing an Olink LOD.

For the 5,420 Olink antibodies (5,416 unique proteins) measured in 102 split samples, the median Spearman correlation was 0.65 (IQI: 0.37-0.90) and the median CV was 35.7% (IQI: 14.5-97.1%; CV<20% for 1,804 assays) (**Figure 1A and Table 1**). The correlations of assays measured in duplicate varied substantially according to the availability of the assay on prior Olink platforms, i.e., Olink 3072 and Olink 96. Assays available on the Olink 96 and Olink 3072 platforms had median correlations of 0.92 (IQI: 0.83, 0.96) and 0.84 (IQI: 0.55, 0.94), respectively, whereas the median correlation for the assays added to the newest Olink Explore HT was 0.45 (IQI: 0.28, 0.66). The corresponding median CVs rose from 10.9% to 19.8% to 67.9% (**Table 1**).

Half of the Olink Explore HT assays had the majority (>50%) of values above the LOD. There were 1,563 assays where all 102 participants had values that were above the published LOD and their median CV was 12.7%. The precision (CV) of the Olink Explore HT assays was strongly inversely correlated (*r*=-0.77) with protein detectability (i.e. percent of samples above the validation LOD) (**Extended Data Figure 1**). When we replaced Olink values reported below the LOD to half the LOD, the median correlation for the Olink assays measured in duplicate increased to 0.79 (IQI: 0.58-0.93), and the median CV decreased to 13.3% (IQI: 8.7-21.7%) (**Figure 1A and Table 1**). Focusing on highly detectable proteins (i.e. with majority of samples above the LOD) left 2,446 assays with a median correlation for duplicate measurements of 0.79 (IQI: 0.59-0.92) and median CV of 13.6% (IQI: 8.9-21.9%). Treating values below the LOD as missing data left 4,743 proteins with a median correlation for duplicate measurements of 0.88 (IQI: 0.68, 0.96) and a median CV of 17.6% (IQI: 11.2%-28.5%).

The between-platform median correlation for the 4,443 overlapping assays on the SomaScan 11k and Olink Explore HT platforms was 0.14 (IQI: 0, 0.58), lower than the 0.33 reported for the earlier versions of these platforms.^1^ The distribution of the correlations for the overlapping assays had a mode of 0 with another smaller peak at *r*∼0.8 (**Figure 1B**). About one-third of the protein pairs had good to excellent cross-platform correlations (407 proteins with *r*≥0.8; 875 proteins with 0.5≤ *r*<0.8) while the other two-thirds had poor correlations (including 2,481 proteins with *r*<0.2). Approximately one-third of the overlapping proteins had 100% of values above the LOD on Olink Explore HT and those assays had a median between-platform correlation of 0.52. Conversely, plasma proteins with the majority of samples below the LOD on the Olink platform showed poor correlation to the corresponding SomaScan assay (**Figure 1B**). For the overlapping assays, the CVs for the Olink Explore HT assays were strongly inversely correlated (*r*=-0.82) with protein detectability (percent of samples above the Olink LOD); the SomaLogic 11k assays were not correlated with protein detectability (r=-0.02).

After replacing protein values reported below the LOD with the LOD divided by 2, the median correlation for the 4,325 proteins on both the SomaScan 11k and Olink Explore HT platforms was 0.13 (IQI: -0.01, 0.58), and these results were similar to our primary analyses (median *r* 0.14).

In conclusion, the field of proteomics is expanding rapidly and thousands of proteins, particularly those in higher abundance, can be measured with great precision. We identified substantial differences in precision within and across the latest large-scale proteomic platforms available. Expanding the coverage of human plasma proteomic profiling retains precision with SomaScan’s aptamer-based technology. In contrast, newly added antibodies on the latest Olink platform often yielded imprecise values below the LOD of the assay in human plasma. Our results provide an important update to the recently published comparison of these leading assays.

## METHODS

### Study Design

The ARIC Study is a community-based prospective cohort study which began in 1987 to 1989 when the participants were aged 45 to 64 years.^2^ The 15,792 participants were recruited from 4 US communities: suburban Minneapolis, Minnesota; Jackson, Mississippi; Forsyth County, North Carolina; Washington County, Maryland. Visit 5 occurred in 2011-2013 and was attended by 6,538 participants (aged 66 to 90). Plasma samples (never previously thawed) were thawed using a standardized quick thaw and refreeze protocol. In 2023, samples were aliquoted and shipped on dry ice to SomaLogic for SomaScan 11k measurements (samples in duplicate) and to Baylor Human Genome Sequencing Center for Olink 5k HT measurements (samples in duplicate).

These analyses are based on data from up to 116 ARIC participants without prevalent coronary heart disease, stroke, or heart failure at visit 5. As previously described,^3^ a random subset of visit 5 participants who developed incident coronary heart disease, stroke, or heart failure within 5 years of visit 5 were selected, and were balanced according to categories of age (≥73 or <73 years), sex, race (Black or White), and eGFR (≥60 or <60 mL/min/1.73m^2^). These participants were frequency matched (1:1) to controls (did not have incident coronary heart disease, stroke, heart failure, or die within 5 years) by age (within 10 years), sex, race and eGFR groupings of cases.

### Aptamer-based Proteomic Platforms

We quantified 11,083 aptamer assays using the SomaScan 11k (v5.0) platform^4^ (SomaLogic; Boulder, Colorado) in plasma from 116 ARIC visit 5 participants using highly multiplexed modified DNA-based aptamer-technology. Briefly, proteins were quantified with relative fluorescence intensity (RFU) that was calibrated and normalized for plate variation (SMP) and using adaptive normalization by maximum likelihood (ANML).^4^ We log_2_ transformed RFU values. SomaLogic does not provide LOD values for assays. One of the 116 participant’s samples from visit 5 did not pass SomaScan 11k QC for ANML SMP; two additional participants had samples which did not pass ARIC QC analyses (protein principal component 1-10 deviated >5 standard deviations).

### Immunoassay Proteomic Platforms

We quantified 5,420 antibody assays using the Olink Explore HT platform^5^ (Olink Proteomics, Uppsala, Sweden) on additional aliquots of plasma from the same ARIC participants using proximity extension assay technology. Briefly, relative protein abundance in plasma was quantified based on the binding of oligonucleotide-labeled antibodies pairs to the target protein. These unique hybridization sequences are then amplified with real-time PCR. Olink proteins were reported on a relative and log_2_ scale as Normalized Protein eXpression (NPX) values. Eleven participants from visit 5 had samples which did not pass Olink Explore HT QC (including a participant whose sample did not pass ARIC QC for SomaScan); one additional participant did not pass ARIC QC analyses (principal component 1-10 deviated >5 standard deviations).

### Measurement of Covariates in ARIC

Participants self-reported their race (Black or White). Body mass index was calculated based on measured height and weight. Diabetes was based on self-report physician diagnosis, glucose-lowering medication use, fasting glucose ≥126 mg/dL or non-fasting glucose ≥200 mg/dL. eGFR was calculated using the 2021 Chronic Kidney Disease Epidemiology equation incorporating creatinine and cystatin-C.^6^ Incident cardiovascular disease was based on a composite of adjudicated events including coronary heart disease, stroke, and heart failure^2^ through 31 Dec 2017.

### Statistical Analysis

Statistical analysis was performed using R version 4.3.0.

#### Within-platform assay precision

Based on split plasma samples, we calculated means (SDs) of the proteins measured in each batch. Among the 102 participants with usable data on both platforms, we then calculated CV (using the Bland-Altman method)^7^ and Spearman’s correlations (*r*) for each assay available on the SomaScan 11k and Olink Explore HT. These statistics for each protein were then summarized using histograms and percentiles. We also examined precision of each protein according to availability of the aptamer on prior SomaScan platforms (based on aptamer seqid on the SomaScan 5k, 7k, or 11k) and availability of the protein on prior Olink platforms (based on uniprotid, OlinkID’s were not transferable across the Olink 96, Explore 3072, or Explore HT platforms).

In our primary analysis, we report results using plate-normalized results and include all NPX protein values as reported to us by Olink (including those below the LOD; intensity normalized values yielded the same overall conclusions). We defined LOD based on the publicly available validation LOD data (https://olink.com/resources-support/document-download-center/ Accessed 15 March 2024). Secondary analyses followed alternative suggested methods (https://olink.com/faq/how-is-the-limit-of-detection-lod-estimated-and-handled/) for dealing with values below the LOD: (1) replacing data below the LOD to a single value (we used the LOD divided by 2), (2) excluding proteins assays with a large proportion of samples below the LOD (we used >50% below LOD), or (3) treating values below the LOD as missing data.

#### Between-platform assay comparisons

There were 102 participants with data available on both the SomaScan 11k and the Olink Explore HT for 4,443 overlapping assays (targeting 3,724 proteins based on uniprotID). We calculated the between-platform correlation for each assay across the 102 individuals. We then summarized the cross-platform correlations (based on measurements from batch 1) in histograms according to categories of protein detectability, i.e., <20%, 20-<50%, 50-<100%, 100% of samples with proteins above the Olink Explore HT LOD values.

## Data Availability

All data produced in the present study are available upon reasonable request to the authors

## Acknowledgements

We thank the ARIC Study staff and participants for their important contributions.

## Author Contributions

Dr. Rooney wrote the first draft of the manuscript and designed the figures. Ms. Chen conducted the data analyses under the supervision of Drs. Rooney and Coresh. All authors provided input on the manuscript.

---

**Extended Data Table 1.**
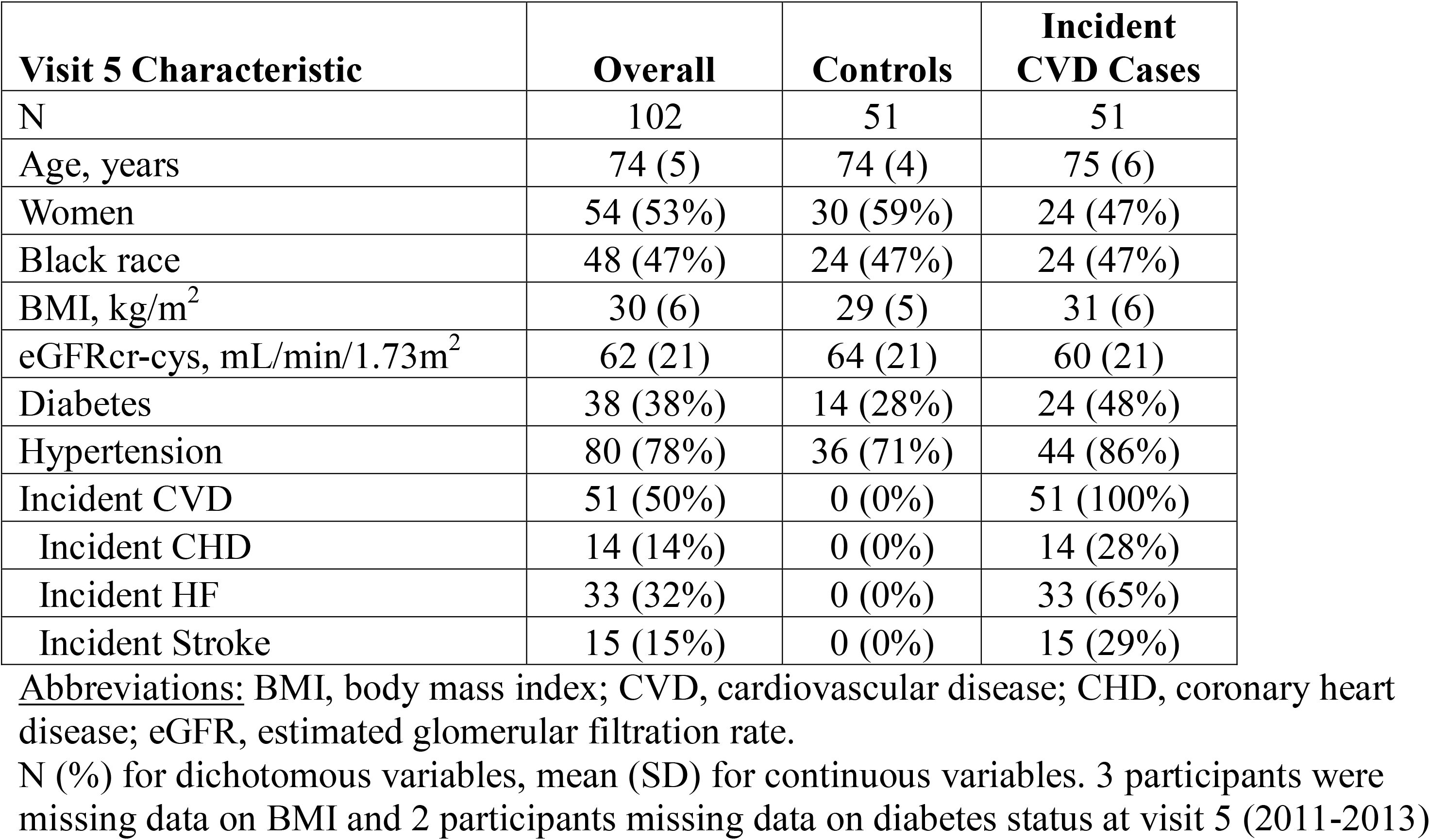
Participant characteristics overall and according to case and control status: The Atherosclerosis Risk in Communities (ARIC) Study (2011-2017).

**Extended Data Figure 1.**
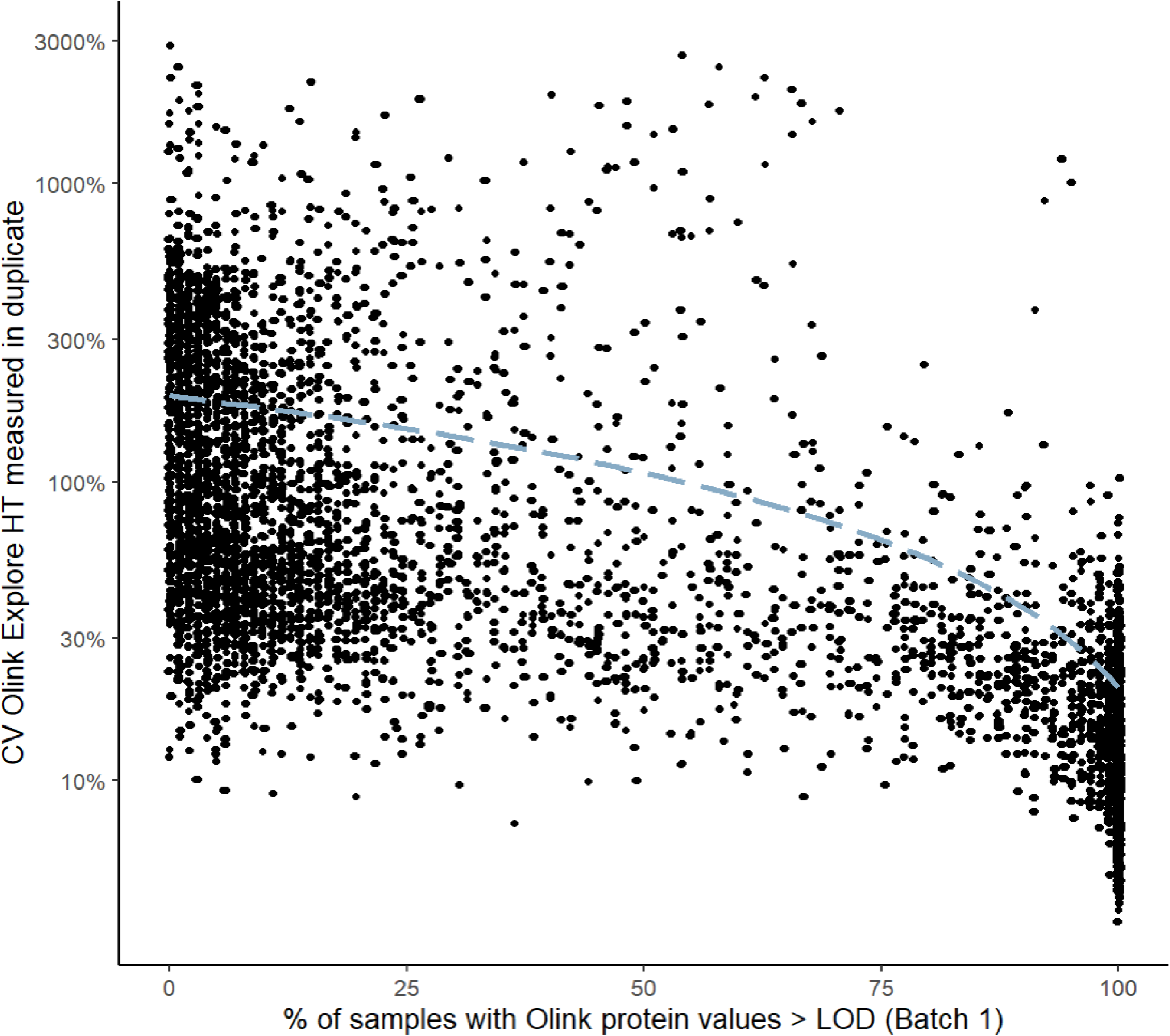
Scatterplot of the percent of participants with protein values above the limit of detection (LOD) versus coefficient of variation (CV, displayed on the log_2_ scale) on the Olink Explore HT among 102 ARIC participants.

